# Follow-up Experiences of Cervical Cancer Patients after Radiotherapy: A Qualitative Study

**DOI:** 10.1101/2025.10.13.25337929

**Authors:** Lan Xiong, Haoyun Wang, Qingcai Wu, Wenwen Cai, Xinyue Deng, Yalan Song

**Affiliations:** Guangzhou Medical University, Guangzhou, Guangdong 510180, China; Affiliated Cancer Hospital of Guangzhou Medical University, Guangzhou, Guangdong 510095, China

## Abstract

**Background:** Cervical cancer remains one of the leading malignancies threatening women’s health worldwide, particularly in developing countries. Radiotherapy is a cornerstone treatment modality but often leads to long-term physical, psychological, and social challenges that significantly impair survivors’ quality of life. Nursing follow-up plays a crucial role in survivorship care; however, patients’ experiences and perspectives during this period remain underexplored in China.This study aimed to explore the subjective experiences of cervical cancer patients during nursing follow-up after radiotherapy, with a particular focus on their perceptions and coping strategies related to physical symptoms, psychological wellbeing, social support, and information needs.

**Methods:** This qualitative study was conducted in the Department of Radiotherapy, Affiliated Cancer Hospital of Guangzhou Medical University, China. Sixteen cervical cancer patients who had completed radiotherapy and entered the follow-up stage were purposively recruited. Guided by supportive care theory, semi-structured in-depth interviews were conducted to explore participants’ experiences.

Data were transcribed verbatim, anonymized, and analyzed using Interpretative Phenomenological Analysis (IPA).

**Results:** The analysis identified six major themes reflecting patients’ multidimensional experiences during nursing follow-up: (i) Persistent discomfort and daily life impact,(ii) Insufficient understanding of the role of nursing follow-up, (iii) Psychological stress and need for support, (iv) Sexual health concerns, (v) Help and conflicts in family support, (vi) Information needs and preferences. These findings highlight both the ongoing difficulties faced by cervical cancer survivors and their expectations for more comprehensive and empathetic follow-up care.

**Conclusion:** Cervical cancer survivors after radiotherapy encounter a complex interplay of physical, psychological, sexual, familial, and informational challenges during follow-up. Patients expressed a strong need for individualized, continuous, and empathetic nursing support to enhance recovery and adaptation. Strengthening the proactivity and professionalism of follow-up services, particularly in sexual health counseling, psychosocial support, and health education, may improve survivorship outcomes. Future multicenter and mixed-methods studies are warranted to inform the development of a more comprehensive follow-up care system.

## Introduction

Cervical cancer is the fourth most common malignancy among women worldwide. Although its incidence and mortality have declined in some countries, it remains highly prevalent in many developing regions ^[1]^. In China, recent statistics (2022) reported 111,820 new cervical cancer cases and 61,579 deaths annually. Cervical cancer ranks sixth in incidence and seventh in mortality among female malignancies, posing a serious threat to women’s health ^[2,3]^.

Radiotherapy (RT) is one of the main treatment modalities for cervical cancer. While effective in eradicating malignant cells, RT inevitably damages surrounding healthy tissues, including the cervix, uterus, vagina, bladder, and rectum ^[4]^. As a result, survivors often experience long-term complications such as pelvic floor dysfunction, urinary and bowel disturbances, and sexual dysfunction, all of which significantly impair quality of life ^[5,6]^. A longitudinal study of cervical cancer survivors found that women who received radiotherapy reported worse physical functioning, somatic symptoms, sexual functioning, and menopausal complaints compared to those treated with surgery alone ^[7]^.

Beyond physical sequelae, cervical cancer survivors face considerable psychological and social challenges ^[8]^. Many women experience uncertainty about fertility after treatment, leading to heightened anxiety and negative emotional responses, which may hinder recovery and coping ^[9]^. The absence of structured psychosocial support during follow-up can exacerbate emotional distress ^[10,11]^. In addition, social stigma associated with gynecological cancers and limited access to reliable health information further compromise survivors’ quality of life. Compared with the general population and survivors of other gynecological malignancies, cervical cancer patients report lower levels of overall life satisfaction ^[12,13]^.

Follow-up care plays a vital role in survivorship management, helping to prevent or mitigate treatment-related physical, psychological, and social issues, thereby improving quality of life ^[14]^. According to the *NCCN Clinical Practice Guidelines in Oncology: Cervical Cancer (Version 4*.*2025)*, recommended follow-up schedules for women after definitive treatment include every 3–4 months in years 1–2, every 6–12 months in years 3–5, and annually after 5 years if no recurrence is detected ^[15]^. However, adherence to follow-up in primary care remains suboptimal, with rates of delayed or missed follow-up ranging from 4% to 75%, particularly among women who are younger, less educated, or socioeconomically disadvantaged ^[16]^.

When nursing follow-up fails to provide systematic and individualized support that addresses both physiological and psychological needs, patients may experience reduced adherence, inadequate information, and increased emotional burdens such as anxiety and distress ^[17]^. A deeper understanding of survivors’ needs and experiences during follow-up requires a multidimensional perspective ^[18]^. Yet, qualitative studies exploring follow-up experiences among cervical cancer patients after radiotherapy remain scarce, especially those capturing patients’ subjective narratives within real-world clinical settings.

To address this gap, this study analyzed interview data from 16 cervical cancer survivors who were undergoing nursing follow-up after radiotherapy. Six major themes were identified: Persistent discomfort and daily life impact, Insufficient understanding of the role of nursing follow-up, Psychological stress and need for support, Sexual health concerns,Help and conflicts in family support, Information needs and preferences.

The following sections present these findings in detail, supported by participant narratives and relevant literature.

## Methods

### Study design and theoretical framework

Guided by the Supportive Care Framework, this study adopted a phenomenological qualitative design to explore women’s subjective experiences during post-radiotherapy nursing follow-up. The approach focused on multi-dimensional needs and responses—physical, psychological, social, informational, and sexual health—and on unmet needs and expectations for care.

### Setting

Fieldwork was conducted from March 1 to June 30, 2025, in the Department of Radiotherapy at the Affiliated Cancer Hospital of Guangzhou Medical University (Guangzhou, China), a regional oncology center with standardized radiotherapy pathways and an established nursing follow-up system. This setting provided a real-world clinical context for the inquiry.

### Participants and recruitment

Participants were women with a confirmed diagnosis of cervical cancer who had completed radiotherapy and entered the follow-up phase (nursing follow-up and/or return visits for clinical review). Purposive sampling was used to recruit patients who could articulate their experiences and consent to participation.

Inclusion criteria: (1) histologically diagnosed cervical cancer; (2) completion of radiotherapy and currently in follow-up; (3) ability to communicate clearly in Mandarin; (4) voluntary participation with written informed consent.

To ensure maximum variation, we considered age, marital status, education, employment, cancer stage, treatment modalities, and time since radiotherapy. Recruitment was supported by ward/clinic nurses through brief announcements and one-to-one screening in inpatient and outpatient areas.

Sixteen women were enrolled; thematic saturation was reached with the 16th interview when no new categories or themes emerged.

### Data collection

A semi-structured interview guide (expert-reviewed and refined after two pilot interviews) covered: understanding of nursing follow-up, bodily and emotional changes, rehabilitation and daily life, social support, and information needs. Trained qualitative researchers conducted all interviews in quiet, private consultation rooms. With permission, interviews were audio-recorded; field notes captured salient non-verbal cues. Each interview lasted ∼30–60 minutes. Clinical charts were reviewed, with consent, to supplement diagnostic, treatment, and follow-up information. Interviews used open-ended questions with probes to elicit depth. The full guide is provided in Appendix 1.

### Data analysis

Interviews were transcribed verbatim and de-identified. Analysis followed Interpretative Phenomenological Analysis (IPA) procedures: iterative reading for immersion; open coding; developing data-driven categories; clustering subthemes; and refining overarching themes supported by exemplar quotations. Coding was facilitated by NVivo 12 (QSR International). Two researchers independently coded transcripts and met to discuss discrepancies until consensus was reached; an audit trail documented analytic decisions.

### Trustworthiness

Credibility was enhanced through brief member checks at the end of each interview (oral summaries for confirmation/clarification). Dependability and confirmability were supported by a detailed protocol and audit trail describing design, instruments, recruitment, data collection, and analysis steps. Transferability was addressed via maximum-variation sampling and thick description with illustrative quotes. To manage bias, the team held regular reflexive meetings to examine assumptions and ensure consistency in theme development.

### Ethics

The study received approval from the hospital ethics committee. All participants received oral and written study information and provided written informed consent, including consent for audio-recording and use of anonymized data for publication. Procedures adhered to the Declaration of Helsinki. Participant confidentiality was protected through coded identifiers and secure data handling.

## Results

### Participant characteristics

A total of sixteen women participated in the in-depth interviews. The mean age of participants was 58.0 years (SD = 12.38), with the majority (62.5%) between 51 and 70 years. Most participants were married, while three were either unmarried or widowed. Educational attainment varied: seven women had a college degree or higher, whereas nine women had secondary education or below, including primary and middle school levels.Regarding occupational status, five participants were employed at the time of interview, three were retired, and the remainder were unemployed or homemakers. Most women resided in urban or peri-urban areas, providing them with basic access to medical facilities.Clinically, eleven participants were diagnosed at stage III cervical cancer. The interval since completion of radiotherapy ranged from 4 months to 8 years, with a mean duration of 22 months. All participants had undergone radiotherapy, with treatment regimens reflecting common clinical practice. Specifically, ten women received combined surgery, chemotherapy, and radiotherapy; three underwent chemotherapy plus radiotherapy; two had surgery plus radiotherapy; and one received surgery plus chemotherapy. This distribution broadly represented the mainstream therapeutic approaches for cervical cancer (Table 1).

**Table 1.** Sociodemographic and Clinical Characteristics of the Participants.

| **Variables** | **Frequencies(n)** | **%** |
| --- | --- | --- |
| **Participants age** |  |  |
| **31–50** | **4** | **25** |
| **51–70** | **10** | **62.5** |
| **71–90** | **2** | **12.5** |
| **Marital status** |  |  |
| **Unmarried** | **3** | **18.75** |
| **Married** | **11** | **68.75** |
| **Widowed** | **1** | **6.25** |
| **Divorced** | **1** | **6.25** |
| **Education level** |  |  |
| **Primary school** | **2** | **12.5** |
| **Middle school** | **6** | **37.5** |
| **High school** | **1** | **6.25** |
| **College and above** | **7** | **43.75** |
| **Occupation** |  |  |
| **Employed** | **5** | **31.25** |
| **Retired** | **3** | **18.75** |
| **Unemployed** | **8** | **50** |
| **Time since radiotherapy ended** |  |  |
| **≤1 year** | **8** | **50** |
| **1–3 years** | **4** | **25** |
| **>3 years** | **4** | **25** |
| **Cancer stage** |  |  |
| **Stage I** | **4** | **25** |
| **Stage II** | **2** | **12.5** |
| **Stage III** | **10** | **62.5** |
| **Treatment method received** |  |  |
| **Surgery + Radiotherapy** | **2** | **12.5** |
| **Chemotherapy + Radiotherapy** | **1** | **6.25** |
| **Surgery + Chemotherapy + Radiotherapy** | **13** | **81.25** |

### Themes identified

Participants reported a wide range of complex experiences across physical, psychological, and social dimensions during nursing follow-up after radiotherapy. The diagnosis and treatment of cervical cancer not only posed challenges to women’s physical health but also had profound impacts on their daily lives, emotional well-being, family relationships, and expectations for the future. From participants’ narratives, six core themes were identified: persistent discomfort and daily life impact, insufficient understanding of the role of nursing follow-up, psychological stress and need for support, sexual health concerns, help and conflicts in family support, and information needs and preferences(Table 2).

**Table 2.** Summary of themes and categories.

| **Themes** | **Categories** |
| --- | --- |
| **(i) Persistent discomfort and daily life impact** | Ongoing physical symptoms and interference |
|  | Pronounced fatigue and functional limitations |
| **(ii) Insufficient understanding of the role of nursing follow-up** | Limited knowledge of nursing follow-up content |
|  | Recognition of the value of follow-up |
| **(iii) Psychological stress and need for support** | Anxiety and depressive emotions |
|  | Low participation in psychological counseling services |
|  | Family and self-adjustment |
| **(iv) Sexual health concerns** | Decline in sexual desire and intercourse difficulties |
|  | Severe lack of sexual health guidance |
|  | Sensitivity and avoidance psychology |
| **(v) Help and conflicts in family support** | Variations in quality of family support |
|  | Recovery of daily functioning and social adaptation |
|  | Coping strategies for economic pressure |
| **(vi) Information needs and preferences** | Preferences in information acquisition methods |
|  | Knowledge gaps |
|  | Views on follow-up format and frequency |

### Theme 1: Persistent discomfort and daily life impact

Most participants reported that they continued to experience varying degrees of physical discomfort after the completion of radiotherapy. These symptoms not only interfered with daily life but also affected psychological well-being and social participation. Such discomfort often persisted or recurred, creating an ongoing burden during the recovery phase. Interview data revealed two subthemes closely related to post-radiotherapy physical symptoms and their impact on daily functioning: Ongoing physical symptoms and interference and Pronounced fatigue and functional limitations.

### Ongoing physical symptoms and interference

Many patients continued to experience bodily symptoms such as lymphedema, urinary and gastrointestinal side effects, and skin discomfort after radiotherapy, which significantly interfered with their daily activities. In addition, the absence of professional rehabilitation guidance led some participants to rely on self-management, potentially increasing health risks.

> “The skin around the radiotherapy site turned dark. I used protective spray, but if it was not applied properly—like the crease between the buttocks—it could easily cause an allergic reaction.” (Participant 3)
>
> “I feel pain when urinating in the morning, especially the first time after waking up. Later in the day, if I drink enough water, the pain subsides. I also feel that radiotherapy has affected my intestines. For example, before treatment I could drink cold water without any issues, but now my intestines feel fragile. Even just a few sips of cold water can trigger diarrhea—it’s as if my intestines have become very sensitive.” (Participant 4)
>
> “My leg often swells up for no clear reason. I think surgery and radiotherapy damaged it, so it keeps swelling and treatment does not help. I usually go for massages outside, but I am not sure whether it is safe. The massage therapist even warned me that improper massage could make it worse.” (Participant 9)

### Pronounced fatigue and functional limitations

Fatigue was a common and distressing problem reported by most interviewees, undermining their independence and work capacity. Some participants also mentioned that fatigue reduced their motivation to go out and engage in social activities.

> “I feel tired so easily… my legs are sore even when sitting. I used to manage some housework, but now I always feel weak. Even a little activity makes me exhausted. Even when resting, my legs feel sore and swollen, and I cannot get relief.” (Participant 2)
>
> “I can’t do anything… my sleep is poor, and I often suffer from insomnia. It takes a long time to fall asleep at night, and I wake up frequently. During the day I already feel weak, and the poor sleep makes it worse. Sometimes I want to do things, but I have no energy, and my body doesn’t cooperate. It makes me feel completely useless.” (Participant 7)

### Theme 2: Insufficient understanding of the role of nursing follow-up

From the interview data, two subthemes were identified regarding patients’ perceptions of nursing follow-up: Limited knowledge of nursing follow-up content and Recognition of the value of follow-up. Overall, although most patients had a preliminary understanding of the term “follow-up,” their comprehension remained limited to the notion of “regular check-ups.” They lacked awareness of the proactive, diverse, and supportive functions of nursing follow-up.

### Limited knowledge of nursing follow-up content

When asked about the meaning of “follow-up,” most participants equated it with “returning on time for examinations.” Many only came back when instructed by physicians and rarely took the initiative to contact nursing staff. Some reported that they had never received a follow-up call from nurses and were uncertain whether they could actively seek consultation. Instead, they mainly relied on medical advice or second-hand experiences shared by others. Such limited understanding of follow-up contrasted with existing clinical guidelines.

> “I thought it just meant coming back on time when the doctor asked me to. When I heard the word ‘follow-up,’ I felt it was simply a task assigned by the doctor—come back to the hospital for a check-up on schedule. I never thought it meant anything else.” (Participant 1)
>
> “I always thought that once radiotherapy was finished, everything was done. Later they told me that I still needed several follow-up visits, and that was the first time I realized it. Actually, I never figured out what follow-up really meant—I only knew I had to come back to the hospital.” (Participant 10)

### Recognition of the value of follow-up

Some participants gradually realized that follow-up was not merely a routine examination but also played a key role in the recovery process. They acknowledged that follow-up could help detect potential problems, remind patients to pay attention to abnormal symptoms, and even allow for timely intervention before conditions worsened.

> “If you detect some warning signs, you can stop it in time. Follow-up is about finding problems earlier, not waiting until the disease becomes serious before dealing with it.” (Participant 3)
>
> “If it hadn’t been for this follow-up discovering the adhesion problem, I might have just kept ignoring it. The nurse asked me some questions and reminded me to get checked, and that was when I realized how serious it was.” (Participant 12)

### Theme 3: Psychological stress and need for support

From the interview data, three subthemes were identified regarding psychological stress and the need for support after radiotherapy: Anxiety and depressive emotions, Low participation in psychological counseling services, and Family and self-adjustment. Overall, the uncertainty associated with the disease and the physical symptoms following treatment placed a psychological burden on most patients. However, their coping strategies varied: some relied on self-adjustment and family support, while others expressed potential needs for professional psychological assistance.

### Anxiety and depressive emotions

Almost all participants mentioned experiencing anxiety, depression, or fear due to the illness and its treatment. These emotional reactions appeared to be stage-specific, with symptoms being particularly prominent in the early period after radiotherapy.

> “Right after completing radiotherapy, I indeed felt anxious, though now I take things more calmly. At that time, I constantly felt fragile, worried about recurrence, and feared that my future life would be heavily affected. At night, lying in bed alone, I felt uneasy and often had racing thoughts. Later, as time passed, I realized my condition was not as bad as I had imagined. I still worry occasionally, but it is not as severe as before, and I can now face it more calmly.” (Participant 8)
>
> “Sometimes I feel scared when I think about this disease, especially when I cannot sleep at night.” (Participant 14)

### Low participation in psychological counseling services

Despite such psychological distress, most participants did not actively seek professional counseling. Some believed they could “think it through” on their own or relied on conversations with family and friends to relieve negative emotions. Only a few joined patient support groups or expressed a desire for regular conversations with nurses.

> “If there were a doctor or nurse who could talk with me regularly, I would feel more reassured.” (Participant 8)

Currently, participation in psychological counseling services remains low, but potential needs do exist. Meanwhile, some patients (such as Participants 4, 10, 11, and 15) reported that their current mental state was stable and that they did not require additional support.

### Family and self-adjustment

In the absence of professional psychological counseling, most patients relied heavily on family members, friends, and self-adjustment strategies to cope with psychological stress.

> “I have support from my family—I have many siblings. They often come to see me, cook for me, and accompany me to follow-up visits. When I am in a bad mood, they comfort me. Although I sometimes worry about my illness, seeing my family around gives me a sense of security, and I do not feel alone.” (Participant 1)

### Theme 4: Sexual health concerns

From the interview data, three subthemes were identified regarding sexual health problems and coping: Decline in sexual desire and intercourse difficulties, Severe lack of sexual health guidance, and Sensitivity and avoidance psychology. Overall, patients commonly experienced diminished sexual function and related physiological problems after radiotherapy. However, due to cultural factors and psychological barriers, sexual health concerns were rarely openly discussed, and systematic professional guidance was lacking.

### Decline in sexual desire and intercourse difficulties

Most participants reported a marked decline in sexual desire after radiotherapy, with sexual activity significantly reduced or even completely discontinued.

> “It’s not that it hurts, but I just don’t feel interested anymore. Life pressures are high, and since the illness and radiotherapy, all my energy has gone into recovery and family matters. Every day I just think about how to get through life. I really have no motivation to think about sex. It’s not that my body is in pain, but I feel it has no meaning, unlike before when I still had desire.” (Participant 1)
>
> “At our age we definitely don’t want it anymore. Besides, I am a patient—I never think about it. And when it’s dry inside, it burns like fire, so painful that I cannot even have the thought of sex.” (Participant 6)

### Severe lack of sexual health guidance

Radiotherapy-related vaginal changes caused clear physical symptoms, especially pain during intercourse. However, most patients did not receive relevant guidance or support. Nearly all emphasized that they had not been given any explanation or instructions on sexual health during follow-up.

> “It definitely hurts when inserted. Without lubricant, it’s impossible. I don’t know the exact time when intercourse is safe. I saw the health education sheet, which said three months, right? But in reality, I didn’t follow it. My ward mate had surgery, and even after more than half a year, she still hasn’t resumed.” (Participant 3) “I asked the doctor, and he said sexual activity is important—it must be maintained, otherwise the endocrine condition gets worse. But regarding how long after radiotherapy it is safe, I am still not sure. I don’t know. We have to wait until the chief physician tells us before we dare.” (Participant 14)

### Sensitivity and avoidance psychology

Participants often expressed feelings of shame, rejection, and self-denial when it came to sexual health, which contributed to these needs being overlooked.

> “I feel that because I am ill, I just don’t think about it anymore.” (Participant 8)
>
> “I feel that since I got sick, I am no longer a normal person.” (Participant 12)
>
> “I feel incomplete now, and psychologically I am somewhat resistant.” (Participant 14)

### Theme 5: Help and conflicts in family support

Family played an important role in the recovery process of patients, but the quality and stability of such support varied considerably. From the interview data, three subthemes were identified: Variations in quality of family support, Recovery of daily functioning and social adaptation, and Coping strategies for economic pressure.

### Variations in quality of family support

Most participants reported receiving care and emotional support from family members, and acknowledged the important role families played in their recovery.

> “My relatives, everyone treats me very well. My husband’s two older sisters care about me, and my son is really considerate. Whenever I need something, he finds a way to help, which makes me feel very warm inside.” (Participant 1)

Some participants, however, noted that although family members were willing to help, they lacked sufficient understanding of post-radiotherapy complications and rehabilitation needs.

> “I feel they may not fully understand my needs. They don’t really know how to help with the aftereffects of radiotherapy.” (Participant 4)

Others described the support as inconsistent, influenced by family members’ own responsibilities and circumstances.

> “My children take turns taking care of me, but they also have their jobs, and I live alone.” (Participant 7)
>
> “My son is tired after work, so he doesn’t pay much attention to me.” (Participant 9)

### Recovery of daily functioning and social adaptation

Some participants reported regaining basic self-care abilities after radiotherapy, such as cooking, sweeping, or light walking. While functional recovery was gradually improving, social adaptation and physical endurance remained limited, highlighting the need for ongoing support and encouragement. A few participants expressed a desire to engage in social activities, but family members’ concerns or their own physical discomfort restricted participation.

> “I don’t go out. Actually, I would really like to, but my family won’t let me. They are afraid I will get too tired.” (Participant 4)
>
> “Now I mainly take care of myself, but when I feel unwell, there are many things I cannot do, and I often need help from others.” (Participant 7)

### Coping strategies for economic pressure

Economic burden was a common concern, with treatment and recovery costs placing significant strain on families. Many participants explicitly mentioned that financial stress was a persistent challenge.

> “Neither of us is working, so of course there is pressure. We rely only on savings and our children’s support, and life always feels tight.” (Participant 5)
>
> “Treatment cost a lot, and I even borrowed money from relatives. Every time I think about repaying the debt, I feel uneasy.” (Participant 7)

### Theme 6: Information needs and preferences

Patients of different ages and levels of information literacy demonstrated distinct preferences in obtaining health information. Their needs varied across information channels, knowledge content, and follow-up models, reflecting the necessity for more individualized and flexible approaches in nursing follow-up. From the interview data, three subthemes were identified: Preferences in information acquisition methods, Knowledge gaps, and Views on follow-up format and frequency.

### Preferences in information acquisition methods

Clear generational and ability-related differences were observed in patients’ preferences for receiving information, indicating that communication strategies should be tailored to patients’ stages and life rhythms. Older patients tended to rely on face-to-face communication, highlighting barriers to using written or electronic media, whereas younger or more digitally literate patients preferred electronic channels.

> “WeChat reminders are very useful. I can check them anytime without making a special trip to the hospital.” (Participant 12)
>
> “Face-to-face communication is better for understanding. Elderly people like me cannot even read. It is best to explain things in person. I cannot use a mobile phone, and I cannot understand phone calls clearly either.” (Participant 15)

### Knowledge gaps

Many participants reported a lack of professional guidance after discharge regarding rehabilitation care, nutrition, and daily management. They not only lacked basic rehabilitation knowledge but also required personalized guidance tailored to their individual conditions.

> “I don’t know much about rehabilitation knowledge. I hope they can teach us some nursing methods, like massage or how to relieve leg swelling.” (Participant 5)
>
> “I am never sure about what I can and cannot eat, especially since I also have diabetes. I need to supplement nutrition but at the same time worry about high blood sugar.” (Participant 2)

### Views on follow-up format and frequency

Participants’ opinions about the frequency and format of follow-up varied depending on their recovery stage and physical condition. Some preferred more flexible and convenient approaches. While a few considered follow-up every three to six months reasonable, others emphasized aligning follow-up with scheduled examinations or using remote methods such as phone or WeChat to reduce unnecessary hospital visits.

> “Right after discharge, I think follow-up should be more frequent. But now, longer intervals are fine.” (Participant 10)
>
> “It’s best to complete follow-up along with examinations, without making an extra trip to the hospital.” (Participant 13)
>
> “Contact by phone or WeChat is also fine—it saves the trouble of going back and forth.” (Participant 16)

## Discussion

This study explored the follow-up experiences and needs of cervical cancer patients after radiotherapy in a tertiary oncology hospital in Guangzhou, and analyzed the gaps in current continuity of care. Participants faced multiple challenges in the physical, psychological, social, and economic domains after treatment. However, variations in functional recovery, psychological adaptation, family support, and economic coping strategies highlighted the necessity of developing individualized nursing follow-up strategies.

Most participants reported persistent physical symptoms after completing radiotherapy for cervical cancer. Lymphedema, urinary and gastrointestinal dysfunction, skin irritation, and pain interfered with daily activities such as household tasks and social participation. These experiences are consistent with previous reports indicating that radiotherapy-induced damage often results in sexual dysfunction, lymphedema, menopausal symptoms, osteoporosis, urogenital and gastrointestinal dysfunction, chronic pain, and fatigue ^[7]^. Although some studies have reported that the overall long-term quality of life among cervical cancer survivors remains acceptable, chronic symptoms and lymphedema significantly compromise quality of life and warrant further attention ^[19]^. In the Chinese context, where patients often return to family and work roles after treatment, these persistent symptoms may further exacerbate limitations in daily living and social engagement. Clinical teams should implement standardized management strategies for common symptoms such as fatigue, including rehabilitation exercises, pain control, and lymphedema care, to improve survivors’ quality of life ^[20,21]^.

The findings also revealed that many patients had insufficient awareness of the importance of structured follow-up and showed low adherence. Studies have demonstrated that cervical cancer survivors often struggle to maintain long-term compliance with regular follow-up visits ^[22]^. On the one hand, some patients mistakenly perceived themselves as “cured” once symptoms were relieved, thereby neglecting the necessity of follow-up. On the other hand, economic burdens, transportation difficulties, and limited access to medical resources further reduced adherence. In this study, some participants explicitly mentioned delays in follow-up due to distance or financial constraints. Within the Chinese cultural context, patients often rely heavily on face-to-face guidance from physicians ^[23]^, and many survivors fail to fully understand the necessity of follow-up plans. Consequently, some only seek medical help when symptoms occur, increasing the risk of delayed detection of recurrence ^[22]^. The latest Chinese clinical guidelines for cervical cancer emphasize the importance of follow-up, requiring regular examinations after radiotherapy ^[24]^. Therefore, clinical practice should strengthen patient education and interventions by providing clear follow-up schedules and content explanations, using reminder systems (e.g., telephone, WeChat), and reducing the burden for patients with financial difficulties.

This study also highlighted that most participants experienced fear of recurrence, emotional distress, and even anxiety or depression during recovery. In China, the prevalence of anxiety and depression among cervical cancer patients has been reported to be as high as 65.6% and 52.2%, respectively ^[25]^. Due to cultural norms, patients often suppress negative emotions to avoid burdening their families, leaving many psychological needs unmet. Previous research showed that approximately 43% of cervical cancer survivors experience clinically significant anxiety or depression during the first three years after treatment, yet fewer than one-quarter of medical centers conduct routine psychological screening ^[22]^. Evidence supports that psychological interventions for cancer survivors, such as cognitive-behavioral therapy and mindfulness-based stress reduction, can significantly reduce fear of recurrence and psychological distress ^[26]^. It is therefore recommended that psychological assessment be integrated into China’s follow-up system, extending mental health services into the community to meet survivors’ long-term emotional and supportive needs.

The impact of cervical cancer treatment on sexual health was profound. Many participants reported decreased sexual desire, vaginal dryness, and dyspareunia, yet often felt too embarrassed to bring up these concerns or received little support^[27]^. More aggressive treatment regimens, such as concurrent chemoradiotherapy with brachytherapy, can cause greater structural and functional damage to the vagina ^[19]^. In this study, some women described feeling distant from their partners. Communication barriers regarding sexual health were striking. Influenced by traditional Chinese values, patients often refrained from discussing sexual issues with physicians, while healthcare providers seldom took the initiative to inquire ^[28,29]^. Clinical follow-up should recognize the importance of sexual health, incorporate it as a regular agenda item, create a private and trusting environment, and encourage patients to express their concerns. Evidence-based interventions should also be provided, such as vaginal dilators, local hormonal therapy, and sexual counseling ^[30]^.

Family played an important role in patient recovery, with many participants describing care and emotional support from spouses, children, and relatives as vital to coping. However, some patients also reported experiencing guilt over imposing financial and emotional burdens on their families. Studies have shown that 65.2% of gynecologic cancer patients have unmet social needs, with family support serving as a key resource for cervical cancer survivors ^[31,32]^. Healthcare professionals should provide regular updates to family members on the patient’s recovery to foster a sense of involvement and sustained motivation for support. Globally, cervical cancer has shown an increasing trend among younger women ^[33,34]^, and in this study, some participants had already returned to work. Therefore, workplace support should also be considered. At the policy level, financial and caregiving assistance for low-income families should be strengthened to reduce social and economic burdens.

The interviews further revealed differing preferences for acquiring health information. Some patients preferred direct face-to-face guidance from physicians, while others actively sought information via the internet and social media. However, patients faced two major challenges in self-directed searches: the complexity of medical terminology and the unreliability of online information ^[23]^. Without simplified patient-friendly materials or guidance toward high-quality sources, survivors are prone to confusion and anxiety, which may undermine adherence and recovery experiences ^[35]^. It is recommended to develop educational materials tailored for cervical cancer survivors (e.g., concise booklets, WeChat articles, online courses) covering follow-up schedules, side-effect management, nutrition and exercise, and fertility concerns. Hospitals could also leverage multimedia platforms, such as official WeChat accounts, to regularly disseminate rehabilitation knowledge and incorporate Q&A sessions into remote follow-up ^[36]^.

## Limitations

This study was a single-center qualitative study with a relatively small sample size, including only 16 cervical cancer patients after radiotherapy from a specialized oncology hospital in Guangzhou. Therefore, the generalizability and representativeness of the findings are limited. As semi-structured interviews and interpretative phenomenological analysis were employed, data interpretation may have been influenced by the researchers’ subjectivity, and bias could not be completely avoided. Furthermore, this study did not conduct in-depth stratified analyses based on patients’ treatment stages or social backgrounds. Future research should expand sample sources and incorporate quantitative methods to further validate the present findings.

## Conclusion

This study provided an in-depth exploration of the subjective experiences of cervical cancer patients during post-radiotherapy nursing follow-up. It revealed their real challenges and needs regarding physical symptoms, perceptions of follow-up, psychological status, sexual health, family support, and information acquisition. The findings suggest that patients generally expect more continuous, individualized, and empathetic follow-up support to enhance rehabilitation quality and social adaptation. Nursing staff should strengthen the proactivity and professionalism of follow-up services, with particular attention to sexual health guidance, psychological support, and health education. Future multi-center and mixed-methods studies are needed to provide evidence for the development of a more comprehensive and effective follow-up system for cervical cancer survivors.

## Data Availability

All relevant data are within the manuscript and its Supporting Information files.

https://www.kdocs.cn/l/clpjRCvhodtk

## Acknowledgments

We sincerely thank the patients who participated in this study and the clinical staff who supported data collection.

## Appendix 1. Semi-Structured Interview Guide

|  |
| --- |
| Interview Questions |
| After completing radiotherapy, have you experienced any physical discomfort? How have these symptoms affected your daily life? Are there any issues that remain unresolved at present? |
| Are you familiar with the concept of “follow-up care”? If yes, how do you understand it? |
| Have healthcare professionals ever conducted nursing follow-up with you? Are you aware of your post-radiotherapy follow-up plan? What are your views on follow-up care? Has the current follow-up been helpful? What problems have you noticed? What additional content do you think should be included? |
| Are you satisfied with the current form of follow-up care? Do you think the timing of follow-up appointments is reasonable? Are there any adjustments needed? What frequency and form of follow-up would you prefer? |
| Under what circumstances would you be unwilling to participate in follow-up care? Under what circumstances would you be willing to attend? |
| After radiotherapy, have you experienced anxiety, depression, or other emotional problems? How have these emotions affected your daily life? Have you received psychological counseling or participated in a support group? If yes, how effective was it? Would you like to receive more mental health support? If so, in what form? |
| After radiotherapy, have you experienced any sexual health problems? Have these issues affected your quality of life? Have you received sexual health guidance? If yes, how effective was it? What kind of support would you like in relation to sexual health? |
| After radiotherapy, have you received adequate family support? Do your family members understand your needs? Have you experienced financial difficulties? |
| Are you familiar with rehabilitation knowledge for cervical cancer patients after radiotherapy? Would you like to receive more guidance about follow-up care? What is your preferred method for obtaining health information? How do you think the healthcare team could improve the provision of such information? |

